# Social risk factors for pediatric asthma exacerbations: A systematic review

**DOI:** 10.1101/2023.09.19.23295732

**Authors:** Shriya Vinjimoor, Caroline Vieira, Colin Rogerson, Arthur Owora, Eneida A Mendonca

## Abstract

**Objective:** This systematic review aims to identify social risk factors that influence pediatric asthma exacerbations.

**Methods:** Cohort studies published between 2010 and 2020 were systematically searched on the OVID Medline, Embase, and PsycInfo databases. Using our established phased inclusion and exclusion criteria, studies that did not address a pediatric population, social risk factors, and asthma exacerbations were excluded. Out of a total of 707 initially retrieved articles, 3 prospective cohort and 6 retrospective cohort studies were included.

**Results:** Upon analysis of our retrieved studies, two overarching domains of social determinants, as defined by Healthy People 2030, were identified as major risk factors for pediatric asthma exacerbations: Social/Community Context and Neighborhood/Built Environment. Social/Community factors including African American race and inadequate caregiver perceptions were associated with increased risk for asthma exacerbations. Patients in high-risk neighborhoods, defined by lower levels of education, housing, and employment, had higher rates of emergency department readmissions and extended duration of stay. Additionally, a synergistic interaction between the two domains was found such that patients with public or no health insurance and residence in high-risk neighborhoods were associated with excess hospital utilization attributable to pediatric asthma exacerbations.

**Conclusion:** Social risk factors play a significant role in influencing the frequency and severity of pediatric asthma exacerbations.

**3-Question Summary Box:** *1. What is the current understanding of this subject?:* The individual impact of social factors such as insurance, neighborhood, and ethnicity on pediatric asthma exacerbations has previously been explored.

*2. What does this report add to the literature?:* This review systematically identifies the relative importance of individual sociodemographic factors and interactions between them. Race, neighborhood risk, insurance status, caregiver perceptions, and a synergistic interaction between health insurance status and neighborhood risk were found to be contributary.

*3. What are the implications for public health practice?:* It is important for providers to educate patients on how their surroundings impact their respiratory health and advocate for increased healthcare access for at-risk populations.

## Introduction

Social factors have a major influence on public health outcomes among children. Asthma is the most prevalent chronic illness among children globally, with around 1 in 12 American children under the age of 18 affected^1^. In 2019 alone, over 44% of pediatric asthma patients reported experiencing one or more acute asthma exacerbations^2^. Although asthma often presents with a relapsing-remitting pattern, the frequency of asthma attacks typically decreases by mid-childhood for most pediatric asthma patients^3^. However, for some patients, acute asthma exacerbations are persistent throughout childhood, resulting in frequent physician visits and medication costs. Asthma patients face serious financial burdens, with an average of over $3,266 of out-of-pocket healthcare expenditures per patient annually. In addition, these patients often face frequent school absences, mental health challenges, and a diminished quality of life^4^.

In a retrospective cohort study by Keet et al. investigating over 1 million children with asthma enrolled on Medicaid, it was found that children residing in inner-city, urban, and poorer neighborhoods, and those with Hispanic and non-Hispanic black ethnicities had a higher risk of asthma-related emergency department visits^5^. Similarly, Harrington et al found that lower parent health literacy was associated with worse pediatric asthma control^6^.

While there is an abundance of such studies evaluating the influence of individual social determinants of health on asthma, little work has been done to synthesize the relative importance of these factors in altering the risk of acute pediatric asthma exacerbations. According to Healthy People 2030, social determinants of health can be synthesized under 5 overarching categories: economic stability, education access and quality, healthcare access and quality, neighborhood and built environment, and social and community context^7^.

This systematic review synthesizes social factors that are associated with the risk of asthma exacerbations among pediatric patients aged 2-18 years and categorizes them based on the Healthy People 2030 classifications.

## Methods

This systematic review follows the 2020 PRISMA Guidelines for Systematic Reviews.

### Search strategy

Development Process: In collaboration with a medical librarian, seven known relevant seed studies were used to identify records within databases. Preliminary search terms were gathered through identifying keywords in the titles and abstracts of the seven seed studies. A pilot search strategy was developed using these keywords and additional search terms were added based on relevance to these first set of terms. The search was limited to English-language only articles. The search strategy was validated by its ability to identify the seven seed studies on the PubMed database. All seven studies were identified.

Final Search Strategy: Databases were systematically searched as of November 2, 2021. The search was limited to English-only manuscripts published between 2010 and 2020. Following a pilot PubMed search, keywords, and MeSH terms for the 3 overarching concepts of asthma, pediatrics, and social factors were searched in the following databases: OVID Medline, Embase, and PsycInfo.

The complete line-by-line search strategy with all searched keywords and MeSH terms in each database is accessible in the Appendix. The retrieved studies were uploaded onto the Covidence software, a tool used for duplicate removal and manual screening of articles based on title, abstract and full text.

### Eligibility Criteria

This study used phased eligibility criteria associated with each step of the screening process. During the database search, limits for English language only articles and articles published between 2010-2020 were added. No restrictions on study design or publication type were instituted at this stage.

### Phase 1: Duplicates removed

Duplicate studies were automatically removed by the Covidence Software.

### Phase 2: Title and abstract screening

Title and abstract screening was manually performed using the Covidence Software. Studies investigating social determinants of health, as categorized by the Healthy People 2030 definition, including race, ethnicity, income, family stability, or neighborhood safety, were included. Any studies looking solely at environmental or physiological risk factors were excluded. Studies not related to asthma, outside the participant age range of 2-18 years of age, full text unavailable/abstract-only papers, non-English manuscripts, studies solely looking at non-social variables (environmental, biological, etc.), and those not published between 2010-2020 were excluded.

### Phase 3: Full text screening

Full text screening was performed manually using the Covidence Software. In addition to meeting all phase 2 criteria, studies investigating the outcome of asthma exacerbations measured as inpatient or outpatient physician visits, ED visits for asthma, and patients with asthma refractory to inhalers or nebulizers were included. Associations were most commonly reported as odds ratios, risk ratios, or frequency of ED visits. Studies with designs other than randomized controlled trials, cohorts, and those investigating ambulatory care of asthma were excluded.

### Selection process

Upon retrieval of studies from the implementation of the search strategy, duplicate articles were automatically excluded by the Covidence software in Phase 1. In Phase 2, two researchers independently reviewed the title and abstract of all remaining studies based on pre-established eligibility criteria. Conflicts were resolved via discussion between the two researchers till a consensus was reached. In Phase 3, full-text screening was performed by two researchers independently and conflicts were resolved in the same manner.

### Data extraction

Articles meeting full text eligibility criteria in Phase 3 went through data extraction. The following factors were collected from each eligible study: Title, Author, Year of Publication, Social Determinant investigated, Outcome measure, Study Design, Statistical Analysis, Setting or study location, Sample Size, Age Group, Data and Data source, Study Aim, Key Results, and Statistical Significance of results.

## Results

### Study selection

Figure 1 shows the process we used to screen and select all included studies for this systematic review. Following the initial database search, 717 studies were retrieved for the systematic review. After 102 duplicates were removed during Phase 1, the remaining 615 studies proceeded to Phase 2. In Phase 2, 566 studies were excluded during the abstract and title screening. Upon resolving conflicts, the remaining 49 studies proceeded to full text screening in Phase 3. During full text review, 40 studies were excluded and 9 proceeded to data extraction. The data sources used by each of our 9 studies varied widely and can be identified on Table 1 [Table 1 near here]. All included studies were cohort studies, with 3 prospective and 6 retrospective designs. Additional study characteristics can be identified on Table 1 and key findings of each study can be found on Table 2 [Table 2 near here].

**Figure 1.**
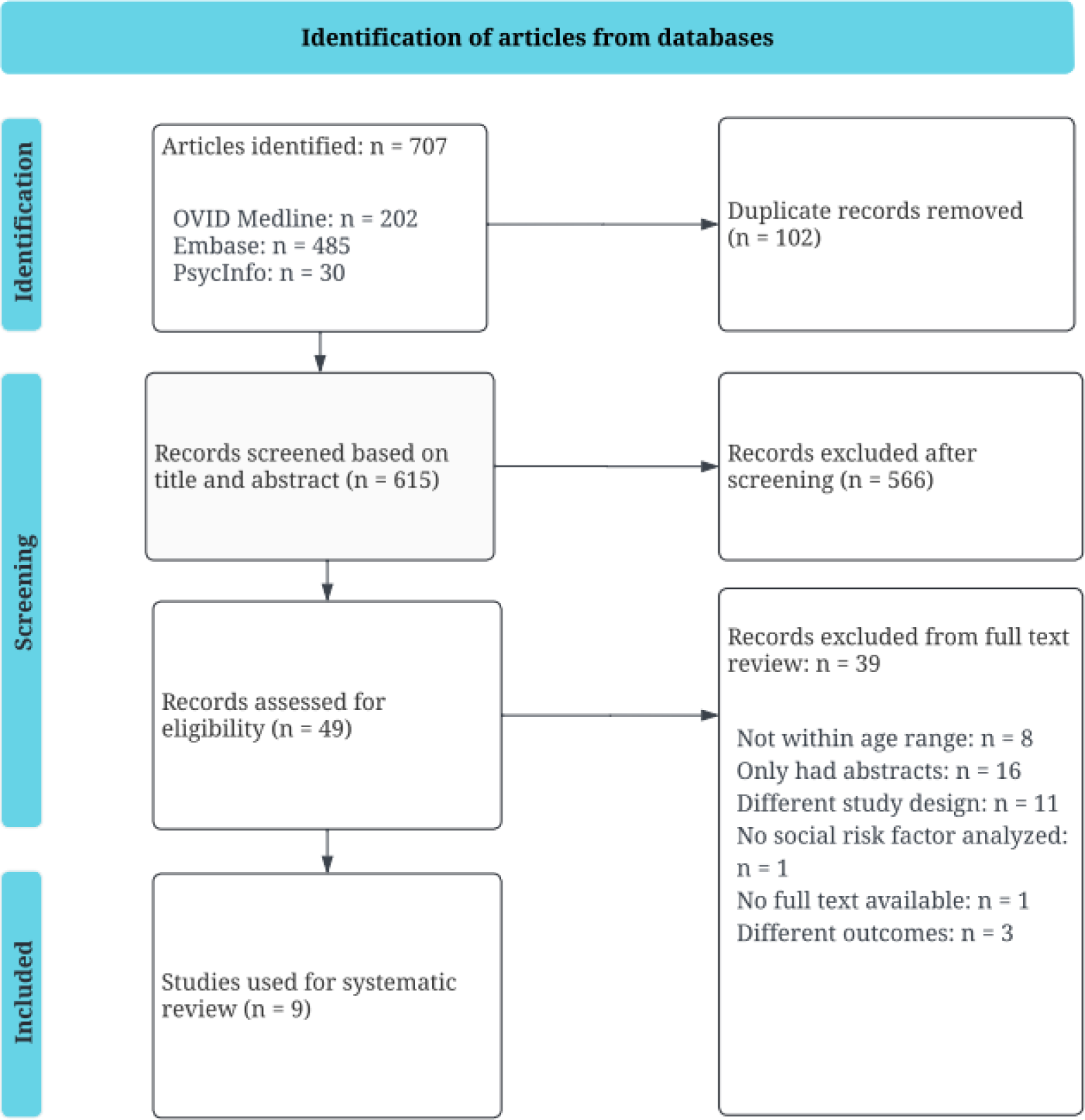
Process of study inclusion and exclusion.

**Table 1:**
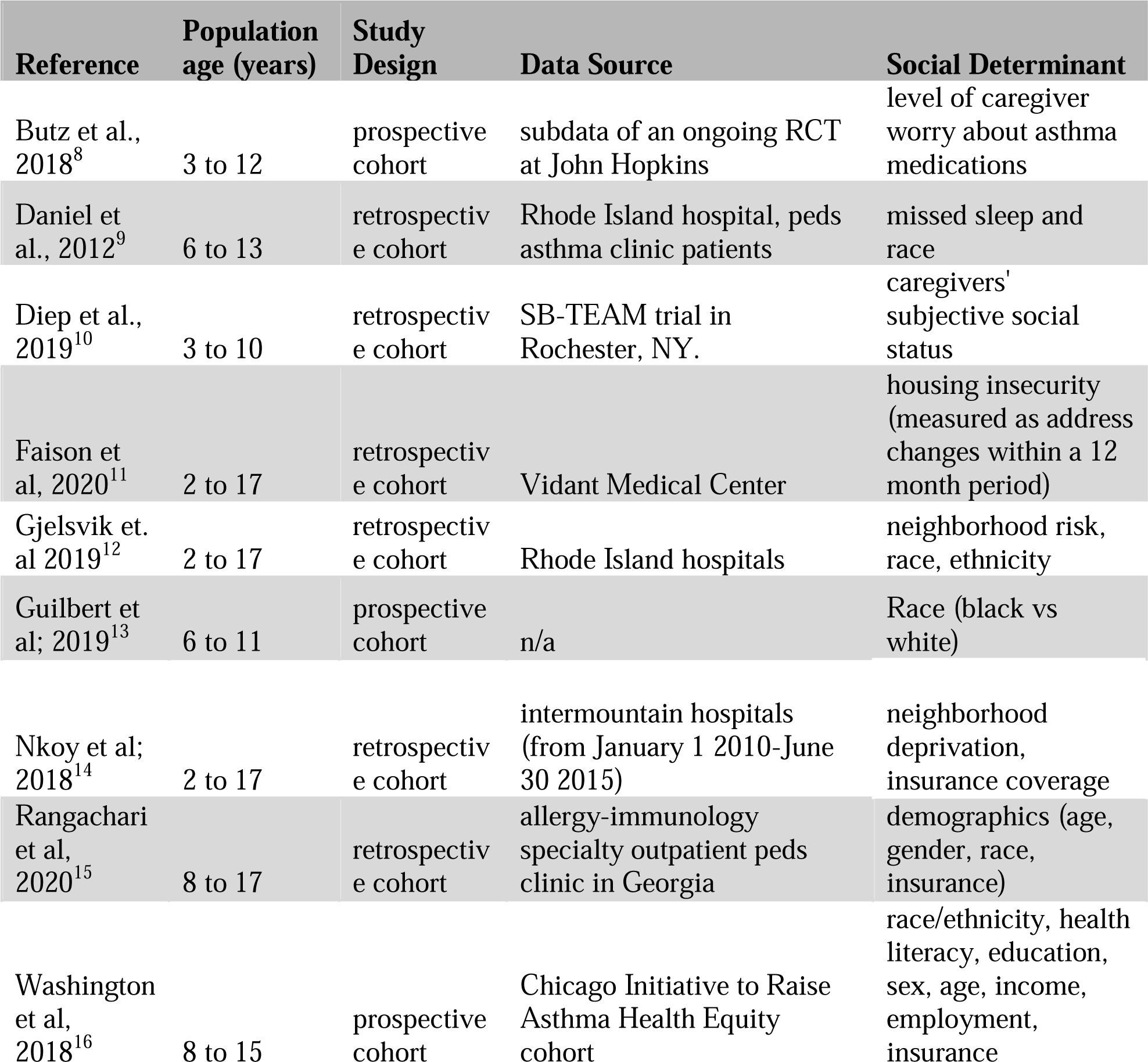
Information from Studies.

| Reference | Population age (years) | Study Design | Data Source | Social Determinant |
| --- | --- | --- | --- | --- |
| Butz et al., 2018<sup>8</sup> | 3 to 12 | prospective cohort | subdata of an ongoing RCT at John Hopkins | level of caregiver worry about asthma medications |
| Daniel et al., 2012<sup>9</sup> | 6 to 13 | retrospective cohort | Rhode Island hospital, peds asthma clinic patients | missed sleep and race |
| Diep et al., 2019<sup>10</sup> | 3 to 10 | retrospective cohort | SB-TEAM trial in Rochester, NY. | caregivers' subjective social status |
| Faison et al, 2020<sup>11</sup> | 2 to 17 | retrospective cohort | Vidant Medical Center | housing insecurity (measured as address changes within a 12 month period) |
| Gjelsvik et. al 2019<sup>12</sup> | 2 to 17 | retrospective cohort | Rhode Island hospitals | neighborhood risk, race, ethnicity |
| Guilbert et al; 2019<sup>13</sup> | 6 to 11 | prospective cohort | n/a | Race (black vs white) |
| Nkoy et al; 2018<sup>14</sup> | 2 to 17 | retrospective cohort | intermountain hospitals (from January 1 2010-June 30 2015) | neighborhood deprivation, insurance coverage |
| Rangachari et al, 2020<sup>15</sup> | 8 to 17 | retrospective cohort | allergy-immunology specialty outpatient peds clinic in Georgia | demographics (age, gender, race, insurance) |
| Washington et al, 2018<sup>16</sup> | 8 to 15 | prospective cohort | Chicago Initiative to Raise Asthma Health Equity cohort | race/ethnicity, health literacy, education, sex, age, income, employment, insurance |

**Table 2:**
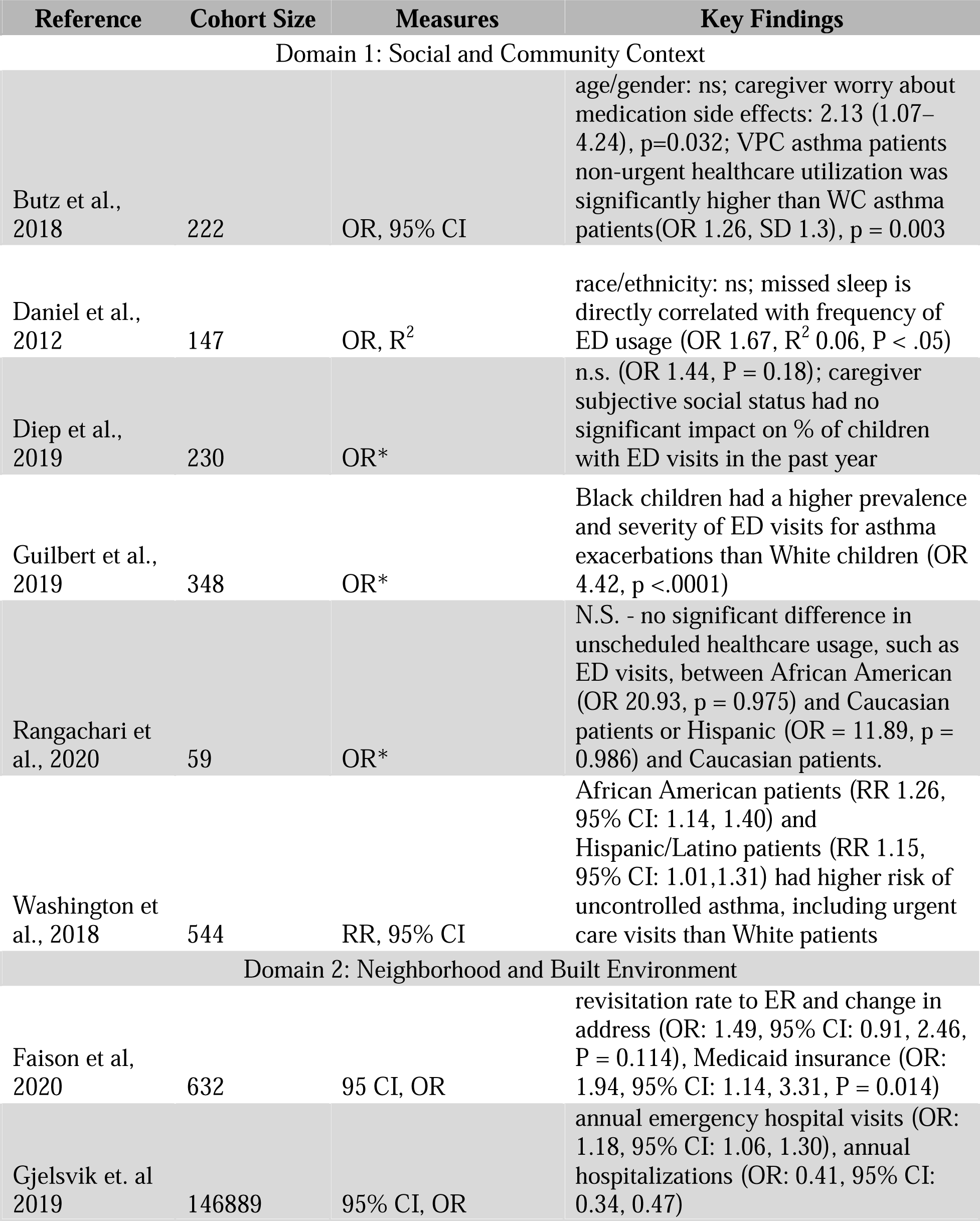
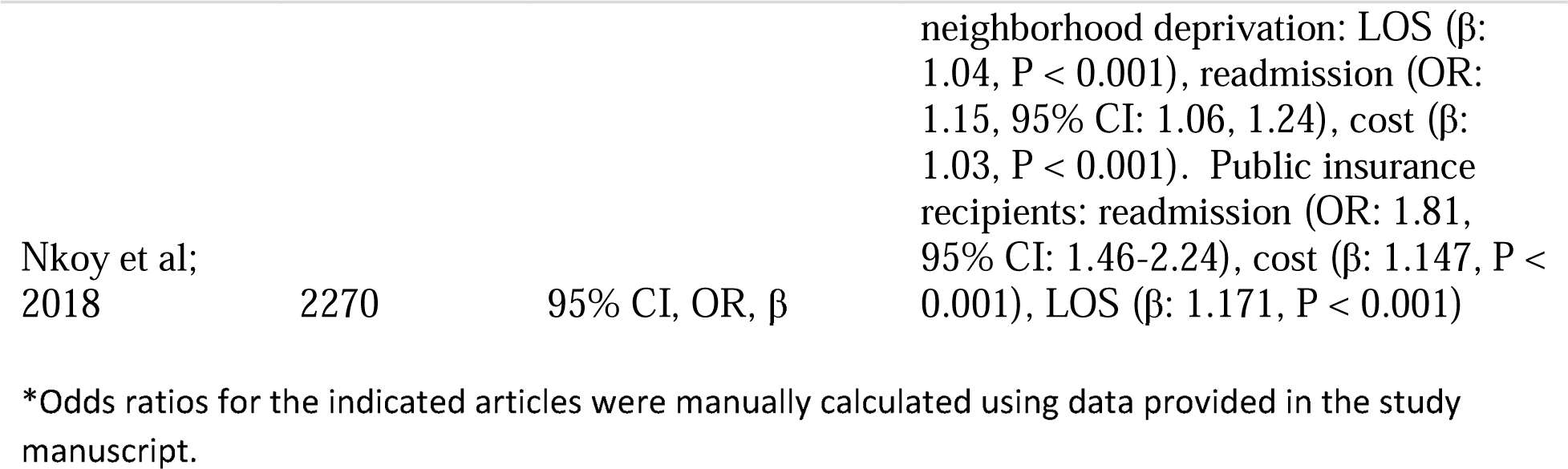
Key Findings from Studies.

### SDoH Domain 1: Social and community context

Six studies explored the role of social and community context in pediatric asthma exacerbations. The influence of three overarching variables were analyzed through the studies retrieved: sociodemographic factors, caregiver perceptions, and patient lifestyle.

### Sociodemographic factors

The influence of sociodemographic factors on pediatric asthma exacerbations has been explored by five studies retrieved from this systematic review: Butz et al., Daniel et al., Guilbert et al., Rangachari et al., and Washington et al.

Three studies found no significant influence of sociodemographic factors on pediatric asthma ED visits. Butz et al conducted a prospective cohort study that collected information on patient demographics and asthma status via caregiver interviews at an initial ED visit and 6 months after. Age and gender were found to have no statistically significant influence on healthcare utilization for very poorly controlled (VPC) asthma, as defined by the NAEPP Guidelines. Similarly, Daniel et al. conducted a retrospective cohort study with self-identified African American, Latino, and Non-Latino White pediatric patients that found no significant association between race and ethnicity and missed sleep and ED visits among pediatric asthma patients when controlling for poverty status. Finally, Rangachari et al. conducted a retrospective cohort study to explore the association between sociodemographic factors and unscheduled healthcare usage for persistent pediatric asthma patients. This study, too, concluded no statistically significant influence of age, gender, and race. Two additional studies found contrary results. Guilbert et al. conducted a prospective cohort study with 6-11 year old asthma patients that either self-identified as Black or White. This study found that Black children were more likely to have an emergency department visit for an asthma exacerbation than White children in the study period (calculated OR 4.42, p <.0001, n = 348). Further, the asthma exacerbations of Black patients were significantly more severe than those of White patients, as measured on the exacerbation hierarchy score (1.2 vs 0.6, p <.001). Similarly, Washington et al conducted a prospective cohort study and found that among 8-15 year-old asthma patients (n = 544), African American patients (RR 1.26, 95% CI: 1.14, 1.40) and Hispanic/Latino patients (RR 1.15, 95% CI: 1.01,1.31) had a significantly higher risk of uncontrolled asthma, measured by the same NAEPP guidelines as Butz et al. This study further found that when controlled for health literacy and education, only African American patients still had a significantly higher risk than White patients for uncontrolled asthma.

### Caregiver perceptions

The influence of caregiver perceptions on pediatric asthma exacerbations was the focus of two studies: Butz et al and Diep et al.

Diep et al conducted a retrospective cohort study using phone interviews on caregivers of 3 to 10 year-old children with persistent asthma. Caregivers’ subjective social status was measured using a self-reported relative perspective from worse off or better off than several groups, including all Americans, their neighbors, and members of their race or ethnicity. The patient’s asthma control was assessed by a number of ED visits for asthma in the past year, a survey of symptoms in the past two weeks, as well as the patient’s score on the standardized Childhood Asthma Control Test. Results indicated no statistically significant difference in the number of pediatric asthma-related ED visits in the past year between high and low caregiver subjective social status (calculated OR 1.44, adjusted *p* = 0.18, n = 230).

The prospective cohort study conducted by Butz et al did find caregiver perceptions to be a significant factor. Caregiver worry about medication side effects increased the odds (OR 2.13, p = 0.032, n = 222) of a patient having very poorly controlled asthma, as diagnosed based on NAEPP guidelines. Consequently, these VPC asthma patients then had increased healthcare utilization compared to patients with well-controlled asthma (OR 1.26, p = 0.003, n = 222), including 1.26 times more frequent primary care visits with no significant difference in ICU admissions. Additionally, around 40% of caregivers of VPC asthma patients rated asthma control as higher than measured by the NAEPP guidelines, showing a disparity between caregivers’ perception of health status and its reality.

### Lifestyle

One retrospective cohort study, by Daniel et al., explored the impact of lifestyle practices. It evaluated the influence of missed sleep on pediatric asthma morbidity, including number of ED visits for asthma as measured on the AFSS asthma morbidity scale. It was found that more frequent missed sleep in the past 12 months was significantly correlated with more frequent ED usage in the past 12 months (OR 1.67, p < .05, n = 147).

### SDoH domain 2: Neighborhood and built environment

Three studies examined the role of neighborhood characteristics and built environment in relation to pediatric asthma. The two main variables pertaining to this domain were as follows: characteristics of neighborhood and effects of change in location.

### Neighborhoods: High and Low Risk

Two studies – Nkoy et al. 2018 and Gjelsvik et al. 2019 – specifically focused on comparing low- and high-risk neighborhoods in relation to the number of hospital visits after an asthma diagnosis.

In Gjelsvik et al., information on patients’ neighborhood risk levels was obtained through data extraction from the 2010-2014 American Community Survey and the 2010 US Census. The study found that children living in the highest poverty risk quintile were more likely to be readmitted to the hospital due to complications of asthma (OR 1.18, 95% CI 1.06,1.30, n = 146,889).

Nkoy et al found similar results. Through data extraction from the 2013 US census block group data and the enterprise data warehouse (EDW), patients’ neighborhood risk was classified using the area of deprivation index (ADI) metric, with high ADIs relating to more deprived populations in terms of education, housing, and employment. It was found that patients residing in higher-risk neighborhoods were more likely to experience higher rates of hospital readmission (OR 1.15, 95% CI 1.06-1.24, n = 2270), along with a longer hospital length of stay compared to their lower-risk counterparts (B = 1.04, p < 0.001, n=2270).

### Change in neighborhoods

Faison et al 2020 focused on exploring if housing instability, assessed as frequent changes in address in the 12 months prior to the study, was associated with increased rates of return ED visits. It was found that patients with at least one address change in a 12-month period were more likely to have severe asthma and more recent visitations. However, address changes were not significantly associated with an increase in return visits to the emergency department for pediatric asthma patients. Moreover, when conducting a multivariate logistical regression of the 90-day revisits, it was found that changes in address were not significantly associated with increased revisitation (OR: 1.49, 95% CI: 0.91, 2.46, p = 0.114, n = 632).

### SDoH Interaction Effects: Insurance and asthma

Several studies investigated the role of patients’ health insurance modality, distinguished as public or private, on pediatric asthma exacerbations. Nkoy et al. 2018, Faison et al. 2020, and Washington et al. 2018 demonstrated a significant association between public health insurance and hospital revisitation due to asthma. Nkoy et al. 2018 and Faison et al. 2020 related this increased rate of revisitation and type of insurance to place of residence, as patients who lived in high-risk areas and possessed public health insurance such as Medicaid were more likely to revisit the hospital due to asthma complications (Nkoy - OR: 1.81; 95% CI:1.46-2.24, n = 2270; Faison - OR: 1.94, 95% CI: 1.14-3.31, p = 0.014, n = 632) than their low-risk neighborhood counterparts. Similarly, Washington et al. 2018 found that patients with non-private health insurance were more likely to experience uncontrolled asthma (RR 1.16, 95% CI 1.08, 1.24, p <.001, n = 544) and lower quality of life (RR –0.33, 95% CI –0.43, -0.23, p <.001, n = 544) than those with private insurance.

## Discussion

This systematic review aimed to identify social risk factors that influence pediatric asthma exacerbations. Overall, our findings suggest that race, neighborhood risk, health insurance status, and caregiver perceptions are important social factors. African American race is associated with an increased risk of asthma exacerbations compared to patients of Caucasian race. Residence in higher-risk neighborhoods was associated with higher ED readmission rates and longer hospital stays compared to lower-risk neighborhoods. Pediatric patients with public or no health insurance were also more prone to higher rates of readmission, longer hospital stays, and higher asthma severity. Finally, caregiver worry about medication side effects and inaccurate perception of patient’s asthma control were associated with higher rates of very poorly controlled asthma.

Reviewed evidence suggests that social/community context and neighborhood/built environment synergistically interact to influence a child’s risk of ED visits for asthma exacerbations. Specifically, race, gender, age, and ethnicity are variables that have been widely explored under social and community context. However, some studies found no significant associations while others identified that Black Race was associated with more frequent and severe asthma exacerbations that needed emergency department care. This finding was supported by previously conducted systematic reviews, including Buelo et al. 2018, in which African American children were found to have an increased risk of experiencing asthma attacks compared to non-African American populations^17^. Further, caregiver perceptions of the patient’s asthma care were found to be significant contributors to asthma exacerbations. This result was echoed in previously conducted reviews, such as Tzeng et al. 2017, in which it was observed that patients whose parents possessed low health literacy were more likely to have poor asthma control^18^. This finding highlights the need for improving caregiver education on asthma management and prognosis by long-term healthcare providers as it could significantly improve patient outcomes.

In exploring the influence of neighborhoods/built environment, it was found that high-risk neighborhoods were correlated with higher lengths of stay and readmission rates for pediatric asthma compared to low-risk, higher-income neighborhoods. However, the frequency of neighborhood change was not significantly associated with an increase in the rate of readmission and length of stay in pediatric asthma patients. These results are similar to those seen in Buelo et al. 2018, which determined that patients with low socioeconomic status had an increased risk for an asthma attack. These findings indicate the need for more resources, such as educational opportunities, public transportation, and access to healthcare resources, to minimize these factors from exacerbating the consequences of asthma further.

Additionally, it was found that the interaction between health insurance type and neighborhood risk had a significant influence on pediatric asthma outcomes. Overall, having public or no health insurance was associated with higher asthma severity and rates of readmission, along with longer hospital stays. These results display the necessity of implementing programs that provide financial support to patients, especially those of lower socioeconomic status.

This study does have limitations. As this topic is not well-studied, a large gap remains in the number of studies available for a review. Several articles were abstract-only publications that did not have a full text associated with them, which is a potential source of publication bias. The study screening was done by two authors. During the title and abstract screening, conflicts were resolved through discussion between these two authors, which could potentially introduce bias. However, the conflicts during full-text screening were resolved by a third reviewer. Additionally, the studies included in this review were limited to the time range of 2010 to 2020, which may limit the generalization of findings to outcomes beyond those years. Despite these limitations, we worked to minimize the potential biases by following the 2020 PRISMA Guidelines for Systematic Reviews.

## Conclusion

Social factors play a role in pediatric asthma. It is vital for healthcare professionals to make patients aware of how their surroundings and lifestyles affect their respiratory health, and to support social movements that enhance the social environments and resources available to at-risk patient populations. Further research using expanded data elements describing social determinants of health for pediatric asthma are needed.

## Data Availability

All data produced in the present work are contained in the manuscript

## Acknowledgements

We are thankful to Mirian Ramirez, from the Ruth Lilly Medical Library, for development of the search strategy. We are also grateful to our lab members, Dr. Eneida Mendonca, Dr. Colin Rogerson, Dr. Arthur Owora, and Dr. Tian He for their support with this project.

## Appendix

### Full Search Strategy

### Database 1: Ovid MEDLINE(R)

Date run: 11/2/2021

Total results: 202

Ovid MEDLINE(R) and Epub Ahead of Print, In-Process, In-Data-Review & Other Non-Indexed Citations, Daily and Versions(R) <1946 to November 01, 2021>

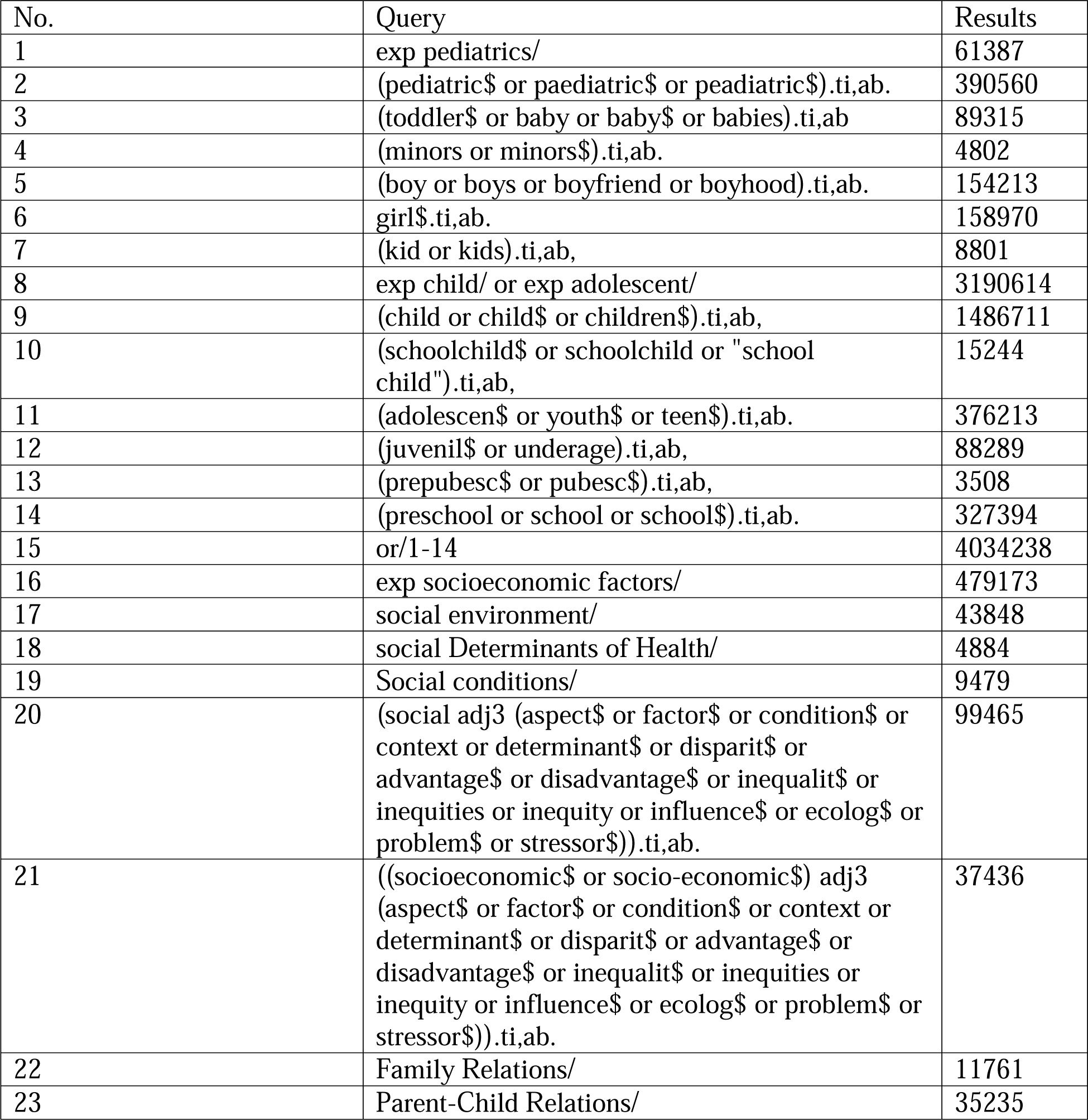

|  |  |  |
| --- | --- | --- |
| 24 | ((family or parent\$ or child\$) adj relat\$).ti,ab. | 15347 |
| 25 | exp "poverty"/ or "poverty".ti,ab. | 64746 |
| 26 | Social Factors/ | 146 |
| 27 | Residence Characteristics/ | 36450 |
| 28 | ((residence or neighborhood\$) adj2 (safety or characteristic\$ or unsaf\$)).ti,ab. | 2258 |
| 29 | "living condition\$".ti,ab. | 8528 |
| 30 | or/16-29 | 684521 |
| 31 | asthma/ | 131647 |
| 32 | Bronchial Hyperreactivity/ | 7471 |
| 33 | respiratory hypersensitivity/ | 9818 |
| 34 | (asthma\$ adj3 (flare-Up or flare-ups or flaring-up or flaring-ups or flareup\$ or exaggerat\$ or acute or magnification\$ or exacerbat\$ or attack\$ or sever\$ or worse\$ or (symptom\$ adj2 increas\$))).ti,ab. | 29787 |
| 35 | (wheez\$ adj3 (flare-Up or flare-ups or flaring-up or flaring-ups or flareup\$ or exaggerat\$ or acute or magnification\$ or exacerbat\$ or attack\$ or sever\$ or worse\$ or (symptom\$ adj2 increas\$))).ti,ab | 1240 |
| 36 | ((bronchial or bronchus or respiratory) adj (hyperreactivity or "hyper reactivity")).ti,ab, | 2358 |
| 37 | (respiratory adj (hypersensitivity or "hyper sensitivity")).ti,ab. | 116 |
| 38 | (airway\$ adj1 (reactive or intermittent or inflammation or hyper-responsiveness or hyperresponsiveness)).ti,ab. | 21060 |
| 39 | 31 or 32 or 33 or 36 or 37 or 38 | 150168 |
| 40 | Symptom Flare Up/ | 901 |
| 41 | (flare-Up or flare-ups or flaring-up or flaring-ups or flareup\$ or exaggerat\$ or acute or magnification\$ or exacerbat\$ or attack\$ or sever\$ or worse\$ or (symptom\$ adj2 increas\$)).ti,ab. | 4502460 |
| 42 | 40 or 41 | 4502762 |
| 43 | 34 or 35 or (39 and 42) | 49833 |
| 44 | 15 and 30 and 43 | 1322 |
| 45 | exp cohort studies/ | 2239611 |
| 46 | cohort\$.tw. | 703689 |
| 47 | (concurrent or follow-up or incidence).tw. | 1886594 |
| 48 | longitudinal.tw. | 277923 |
| 49 | retrospective\$.tw. | 864272 |
| 50 | prospective\$.tw. | 774379 |
| 51 | or/45-50 | 4086049 |

|  |  |  |
| --- | --- | --- |
| 52 | 44 and 51 | 505 |
| 53 | limit 52 to yr=2010-2020 | 207 |
| 54 | 53 and english.la. | 202 |

### Database 2: EMBASE

Date run:11/2/2021

Total results: 485

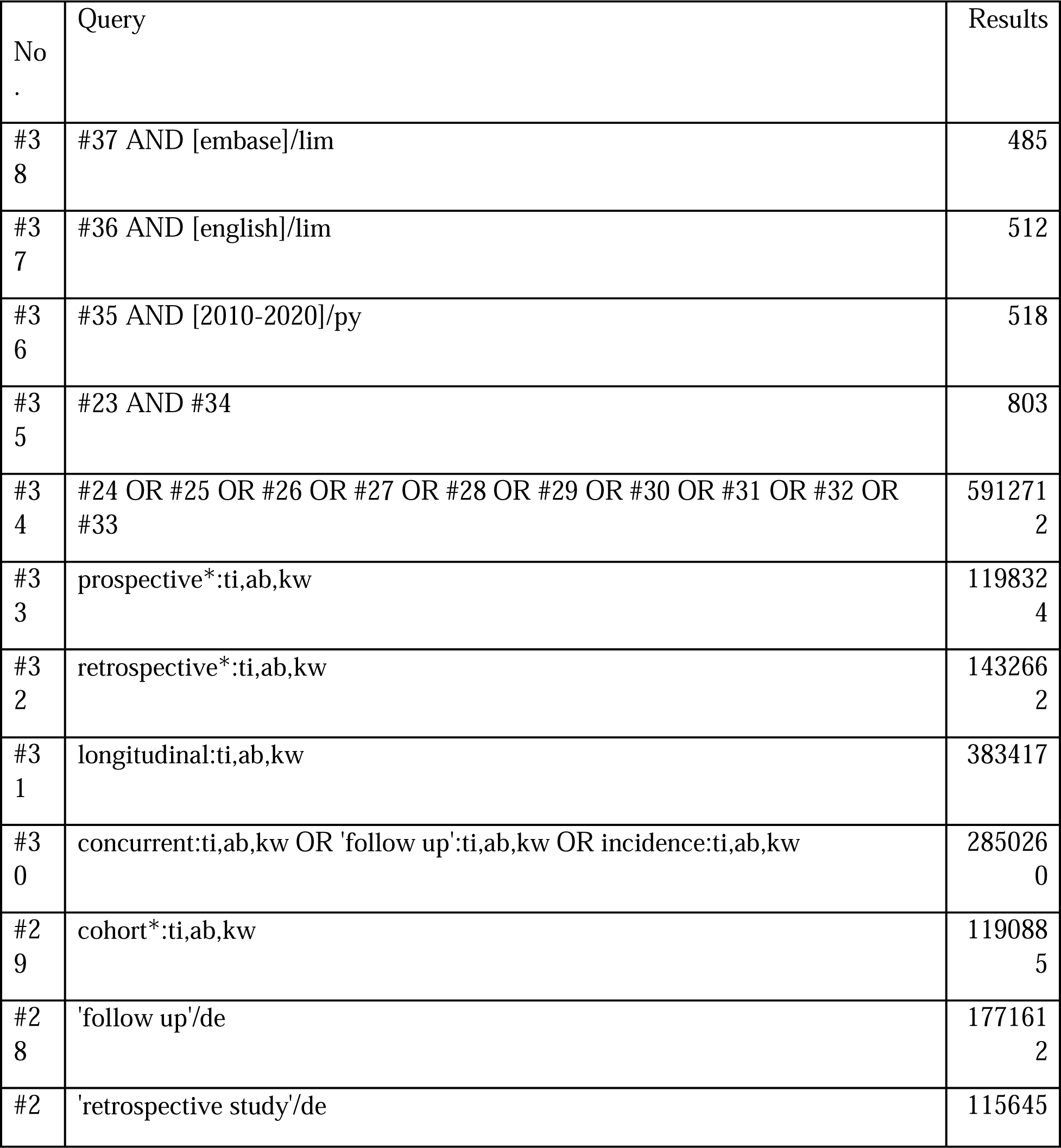

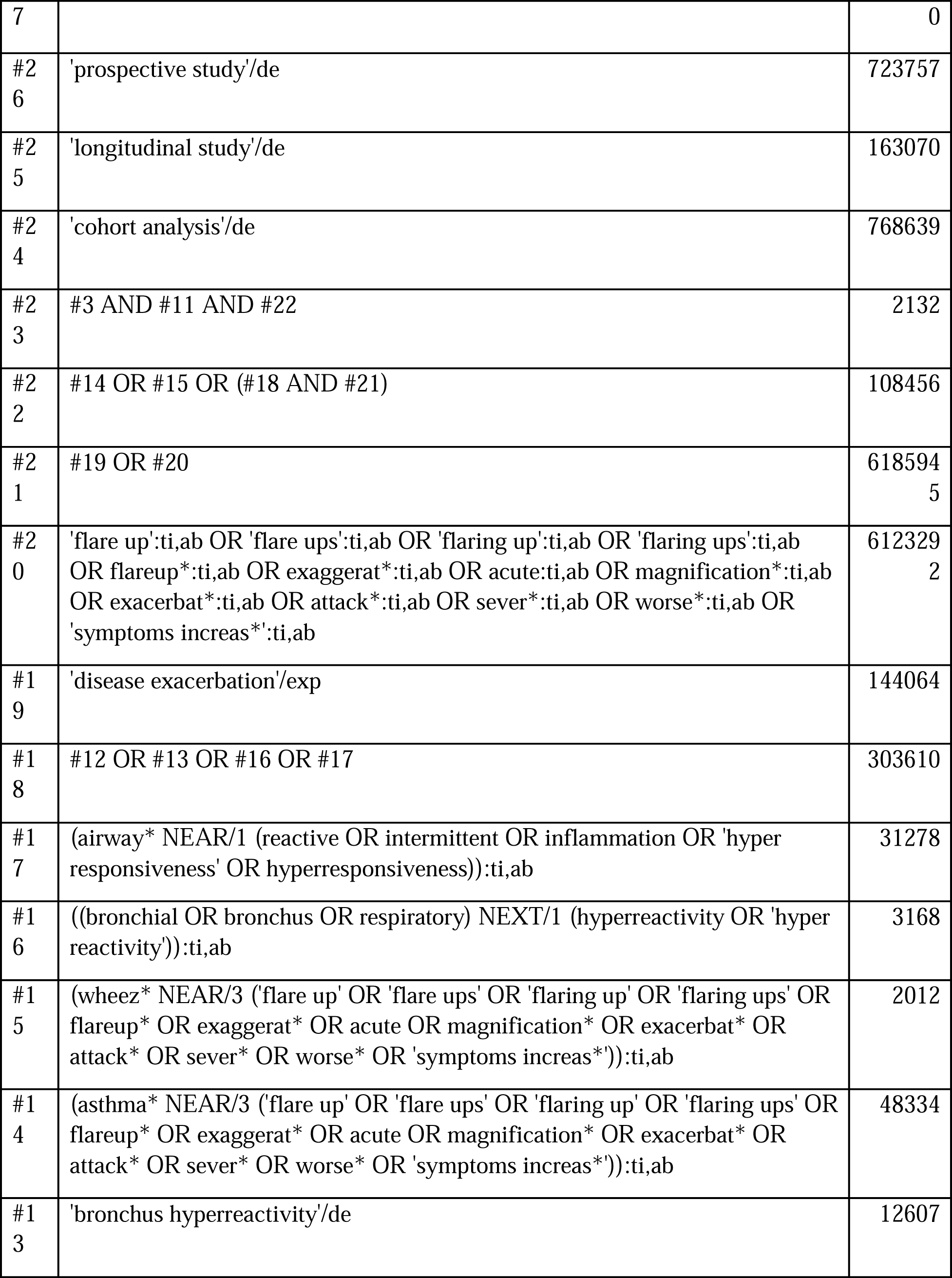

|  |  |  |
| --- | --- | --- |
| #1<br>2 | 'asthma'/exp | 288992 |
| #1<br>1 | #4 OR #5 OR #6 OR #7 OR #8 OR #9 OR #10 | 851990 |
| #1<br>0 | 'living condition*':ti,ab | 10166 |
| #9 | ((residence OR neighborhood*) NEAR/2 (safety OR characteristic* OR unsaf*)):ti,ab | 2569 |
| #8 | 'poverty'/de OR 'poverty':ti,ab | 62941 |
| #7 | 'family relation'/exp OR (((family OR parent* OR child*) NEXT/1 relat*)):ti,ab) | 127164 |
| #6 | ((socioeconomic* OR 'socio economic*') NEAR/3 (aspect* OR factor* OR condition* OR context OR determinant* OR disparit* OR advantage* OR disadvantage* OR inequalit* OR inequities OR inequity OR influence* OR ecolog* OR problem* OR stressor*)):ti,ab | 46027 |
| #5 | (social NEAR/3 (aspect* OR factor* OR condition* OR context OR determinant* OR disparit* OR advantage* OR disadvantage* OR inequalit* OR inequities OR inequity OR influence* OR ecolog* OR problem* OR stressor*)):ti,ab | 121719 |
| #4 | 'social aspects and related phenomena'/de OR 'social aspect'/de OR 'social environment'/de OR 'social status'/de OR 'social problem'/de OR 'social condition'/de OR 'social determinants of health'/de OR 'socioeconomic factors'/exp | 645889 |
| #3 | #1 OR #2 | 490752<br>9 |
| #2 | pediatric*:ti,ab OR paediatric*:ti,ab OR peadiatric*:ti,ab OR toddler*:ti,ab OR baby:ti,ab OR baby*:ti,ab OR babies:ti,ab OR minors:ti,ab OR minors*:ti,ab OR boy:ti,ab OR boys:ti,ab OR boyfriend:ti,ab OR boyhood:ti,ab OR girl*:ti,ab OR kid:ti,ab OR kids:ti,ab OR child:ti,ab OR child*:ti,ab OR children*:ti,ab OR schoolchild*:ti,ab OR adolescen*:ti,ab OR youth*:ti,ab OR teen*:ti,ab OR juvenil*:ti,ab OR underage:ti,ab OR prepubesc*:ti,ab OR pubesc*:ti,ab OR preschool:ti,ab OR school:ti,ab OR school*:ti,ab | 296752<br>7 |
| #1 | 'pediatrics'/de OR 'adolescent'/de OR 'child'/exp OR 'childhood'/de OR 'juvenile'/de | 404384<br>6 |

### Database 3: PsycINFO 1887-current (EBSCO)

Date run: 11/2/2021

total results: 30

SOCIAL RISK FACTORS FOR PEDIATRIC ASTHMA EXACERBATIONS
| # | Query | Limiters/Expanders | Last Run Via | Results |
| --- | --- | --- | --- | --- |
| S46 | (S37 AND S44) | Limiters -<br>Publication Year:<br>2010-2020<br>Expanders - Apply<br>equivalent subjects<br>Search modes -<br>Boolean/Phrase | Interface -<br>EBSCOhost Research<br>Databases<br>Search<br>Screen -<br>Advanced<br>Search<br>Database -<br>APA<br>PsycInfo | 30 |
| S45 | (S37 AND S44) | Expanders - Apply<br>equivalent subjects<br>Search modes -<br>Boolean/Phrase | Interface -<br>EBSCOhost Research<br>Databases<br>Search<br>Screen -<br>Advanced<br>Search<br>Database -<br>APA<br>PsycInfo | 54 |
| S44 | (S38 OR S39 OR S40 OR S41 OR S42 OR S43) | Expanders - Apply<br>equivalent subjects<br>Search modes -<br>Boolean/Phrase | Interface -<br>EBSCOhost Research<br>Databases<br>Search<br>Screen -<br>Advanced<br>Search<br>Database -<br>APA | 472,432 |

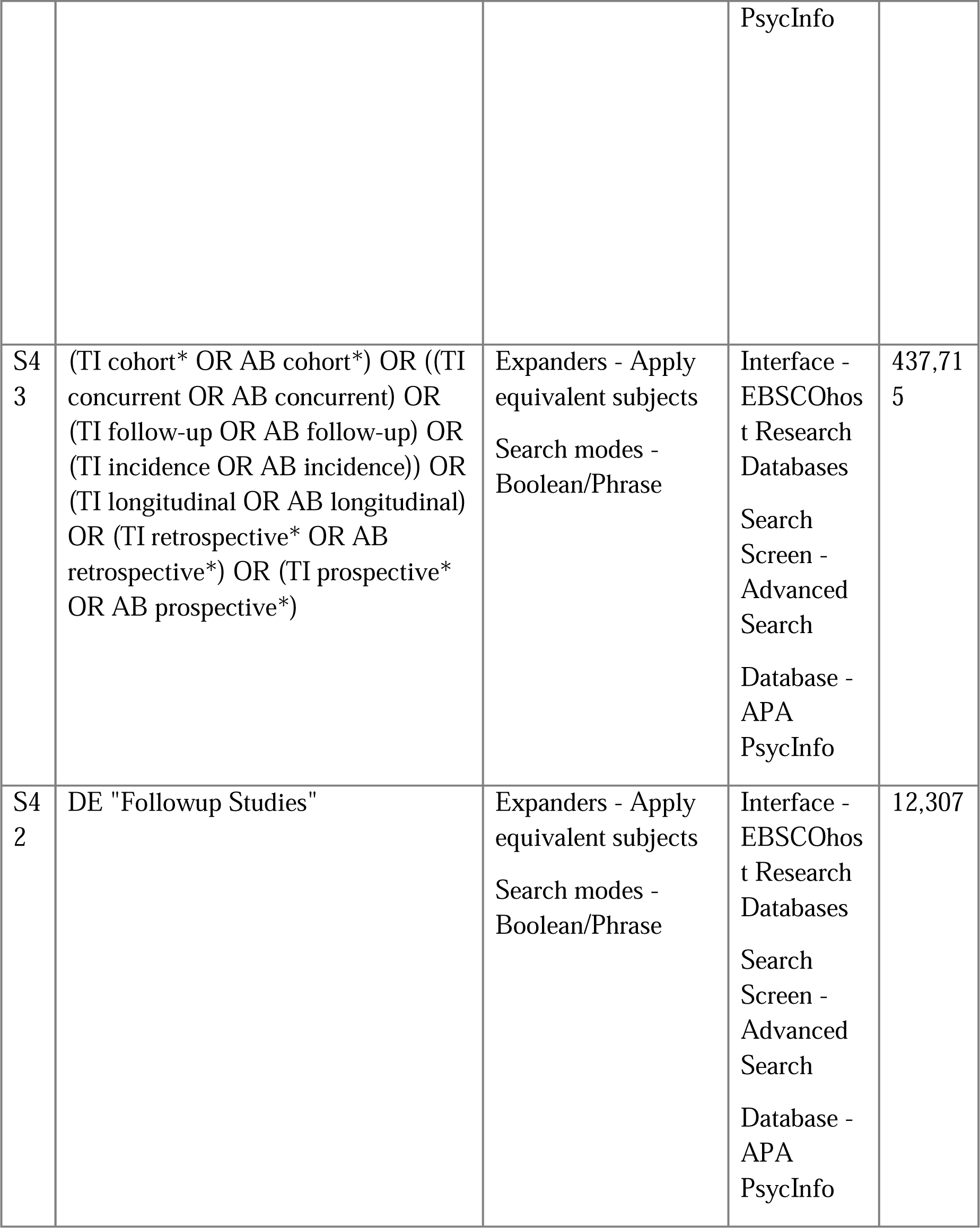

|  |  |  |  |  |
| --- | --- | --- | --- | --- |
| S4<br>1 | DE "Retrospective Studies" | Expanders - Apply<br>equivalent subjects<br>Search modes -<br>Boolean/Phrase | Interface -<br>EBSCOhost Research<br>Databases<br>Search<br>Screen -<br>Advanced<br>Search<br>Database -<br>APA<br>PsycInfo | 28,812 |
| S4<br>0 | DE "Prospective Studies" | Expanders - Apply<br>equivalent subjects<br>Search modes -<br>Boolean/Phrase | Interface -<br>EBSCOhost Research<br>Databases<br>Search<br>Screen -<br>Advanced<br>Search<br>Database -<br>APA<br>PsycInfo | 33,468 |
| S3<br>9 | DE "Longitudinal Studies" | Expanders - Apply<br>equivalent subjects<br>Search modes -<br>Boolean/Phrase | Interface -<br>EBSCOhost Research<br>Databases<br>Search<br>Screen -<br>Advanced<br>Search<br>Database -<br>APA<br>PsycInfo | 51,967 |

|  |  |  |  |  |
| --- | --- | --- | --- | --- |
| S3<br>8 | DE "Cohort Analysis" | Expanders - Apply<br>equivalent subjects<br>Search modes -<br>Boolean/Phrase | Interface -<br>EBSCOhost Research<br>Databases<br>Search<br>Screen -<br>Advanced<br>Search<br>Database -<br>APA<br>PsycInfo | 1,557 |
| S3<br>7 | S7 AND S26 AND S36 | Expanders - Apply<br>equivalent subjects<br>Search modes -<br>Boolean/Phrase | Interface -<br>EBSCOhost Research<br>Databases<br>Search<br>Screen -<br>Advanced<br>Search<br>Database -<br>APA<br>PsycInfo | 262 |
| S3<br>6 | S28 OR S29 OR (S32 AND S34) | Expanders - Apply<br>equivalent subjects<br>Search modes -<br>Boolean/Phrase | Interface -<br>EBSCOhost Research<br>Databases<br>Search<br>Screen -<br>Advanced<br>Search<br>Database -<br>APA<br>PsycInfo | 2,191 |

|  |  |  |  |  |
| --- | --- | --- | --- | --- |
| S3<br>5 | S33 OR S34 | Expanders - Apply<br>equivalent subjects<br>Search modes -<br>Boolean/Phrase | Interface -<br>EBSCOhost Research<br>Databases<br>Search<br>Screen -<br>Advanced<br>Search<br>Database -<br>APA<br>PsycInfo | 668,771 |
| S3<br>4 | (TI asthma* N3 (TI flare-Up OR TI<br>flare-ups OR TI flaring-up OR TI<br>flaring-ups OR TI flareup* OR TI<br>exaggerat* OR TI acute OR TI<br>magnification* OR TI exacerbat* OR<br>TI attack* OR TI sever* OR TI<br>worse* OR TI "symptom increas*"))<br>OR (AB flare-Up OR AB flare-ups<br>OR AB flaring-up OR AB flaring-ups<br>OR AB flareup* OR AB exaggerat*<br>OR AB acute OR AB magnification*<br>OR AB exacerbat* OR AB attack*<br>OR AB sever* OR AB worse* OR<br>AB "symptom increas*")) | Expanders - Apply<br>equivalent subjects<br>Search modes -<br>Boolean/Phrase | Interface -<br>EBSCOhost Research<br>Databases<br>Search<br>Screen -<br>Advanced<br>Search<br>Database -<br>APA<br>PsycInfo | 665,700 |
| S3<br>3 | DE "Severity (Disorders)" | Expanders - Apply<br>equivalent subjects<br>Search modes -<br>Boolean/Phrase | Interface -<br>EBSCOhost Research<br>Databases<br>Search<br>Screen -<br>Advanced<br>Search<br>Database -<br>APA<br>PsycInfo | 19,841 |

|  |  |  |  |  |
| --- | --- | --- | --- | --- |
| S3<br>2 | S27 OR S30 OR S31 | Expanders - Apply<br>equivalent subjects<br>Search modes -<br>Boolean/Phrase | Interface -<br>EBSCOhost Research<br>Databases<br>Search<br>Screen -<br>Advanced<br>Search<br>Database -<br>APA<br>PsycInfo | 5,617 |
| S3<br>1 | ((TI airway* OR AB airway*) N1<br>((TI reactive OR AB reactive) OR (TI<br>intermittent OR AB intermittent) OR<br>(TI inflammation OR AB<br>inflammation) OR (TI "hyper<br>responsiveness" OR AB "hyper<br>responsiveness") OR (TI<br>hyperresponsiveness OR AB<br>hyperresponsiveness))) | Expanders - Apply<br>equivalent subjects<br>Search modes -<br>Boolean/Phrase | Interface -<br>EBSCOhost Research<br>Databases<br>Search<br>Screen -<br>Advanced<br>Search<br>Database -<br>APA<br>PsycInfo | 168 |
| S3<br>0 | ((((TI bronchial OR AB bronchial)<br>OR (TI bronchus OR AB bronchus)<br>OR (TI respiratory OR AB<br>respiratory)) N1 ((TI hyperreactivity<br>OR AB hyperreactivity) OR (TI<br>"hyper reactivity" OR AB "hyper<br>reactivity")))) | Expanders - Apply<br>equivalent subjects<br>Search modes -<br>Boolean/Phrase | Interface -<br>EBSCOhost Research<br>Databases<br>Search<br>Screen -<br>Advanced<br>Search<br>Database -<br>APA<br>PsycInfo | 13 |

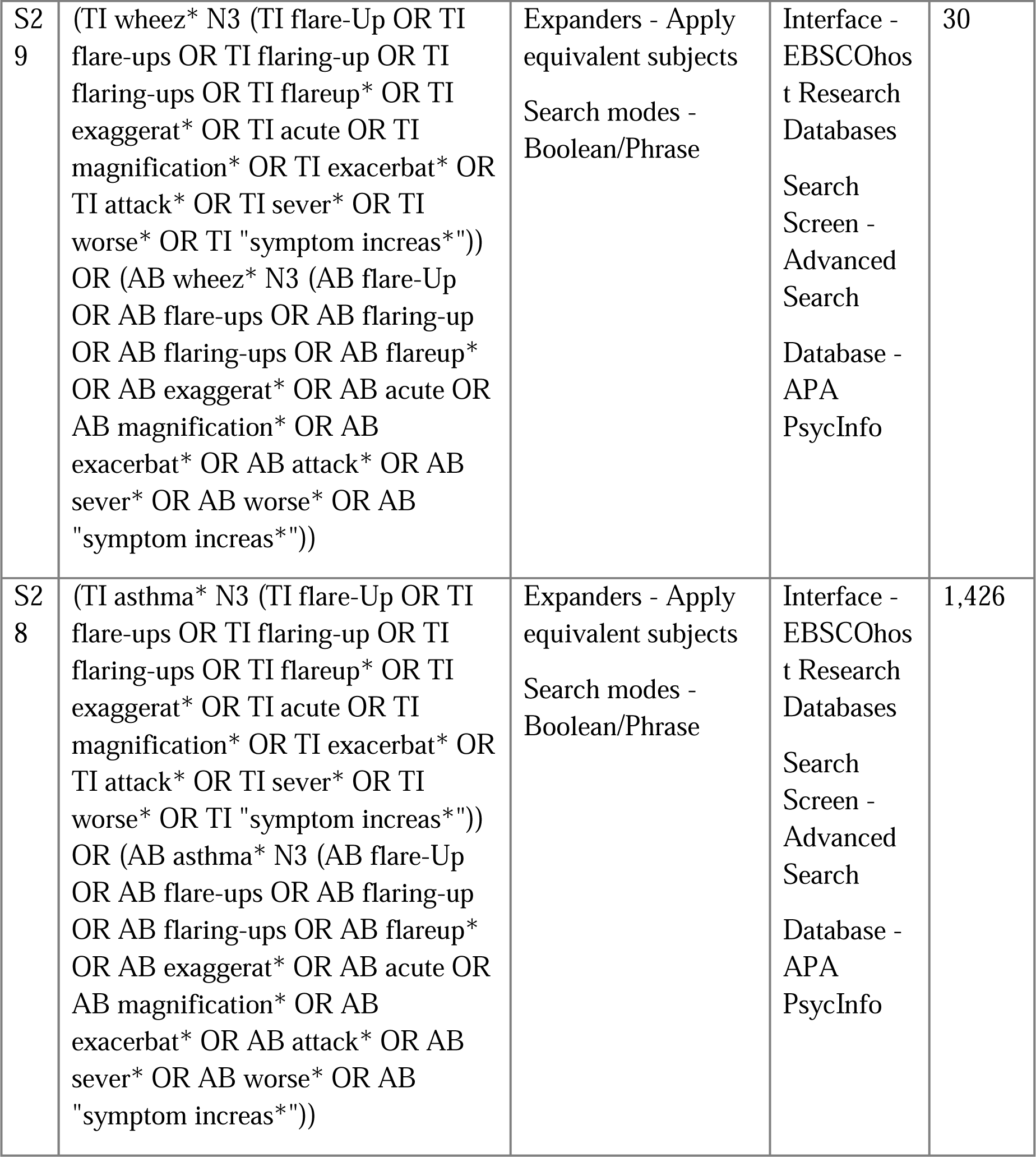

|  |  |  |  |  |
| --- | --- | --- | --- | --- |
| S2<br>9 | (TI wheez* N3 (TI flare-Up OR TI flare-ups OR TI flaring-up OR TI flaring-ups OR TI flareup* OR TI exaggerat* OR TI acute OR TI magnification* OR TI exacerbat* OR TI attack* OR TI sever* OR TI worse* OR TI "symptom increas*"))<br>OR (AB wheez* N3 (AB flare-Up OR AB flare-ups OR AB flaring-up OR AB flaring-ups OR AB flareup* OR AB exaggerat* OR AB acute OR AB magnification* OR AB exacerbat* OR AB attack* OR AB sever* OR AB worse* OR AB "symptom increas*")) | Expanders - Apply equivalent subjects<br>Search modes - Boolean/Phrase | Interface - EBSCOhost Research Databases<br>Search Screen - Advanced Search<br>Database - APA PsycInfo | 30 |
| S2<br>8 | (TI asthma* N3 (TI flare-Up OR TI flare-ups OR TI flaring-up OR TI flaring-ups OR TI flareup* OR TI exaggerat* OR TI acute OR TI magnification* OR TI exacerbat* OR TI attack* OR TI sever* OR TI worse* OR TI "symptom increas*"))<br>OR (AB asthma* N3 (AB flare-Up OR AB flare-ups OR AB flaring-up OR AB flaring-ups OR AB flareup* OR AB exaggerat* OR AB acute OR AB magnification* OR AB exacerbat* OR AB attack* OR AB sever* OR AB worse* OR AB "symptom increas*")) | Expanders - Apply equivalent subjects<br>Search modes - Boolean/Phrase | Interface - EBSCOhost Research Databases<br>Search Screen - Advanced Search<br>Database - APA PsycInfo | 1,426 |

|  |  |  |  |  |
| --- | --- | --- | --- | --- |
| S2<br>7 | (DE "Asthma") | Expanders - Apply<br>equivalent subjects<br>Search modes -<br>Boolean/Phrase | Interface -<br>EBSCOhost Research<br>Databases<br>Search<br>Screen -<br>Advanced<br>Search<br>Database -<br>APA<br>PsycInfo | 5,530 |
| S2<br>6 | (S8 OR S9 OR S10 OR S11 OR S12<br>OR S13 OR S14 OR S15 OR S16 OR<br>S17 OR S18 OR S19 OR S20 OR<br>S22 OR S23 OR S24 OR S25 ) | Expanders - Apply<br>equivalent subjects<br>Search modes -<br>Boolean/Phrase | Interface -<br>EBSCOhost Research<br>Databases<br>Search<br>Screen -<br>Basic<br>Search<br>Database -<br>APA<br>PsycInfo | Display |
| S2<br>5 | (TI "living condition*" OR AB<br>"living condition*") | Expanders - Apply<br>equivalent subjects<br>Search modes -<br>Boolean/Phrase | Interface -<br>EBSCOhost Research<br>Databases<br>Search<br>Screen -<br>Basic<br>Search<br>Database -<br>APA<br>PsycInfo | Display |

|  |  |  |  |  |
| --- | --- | --- | --- | --- |
| S2<br>4 | (((TI residence OR AB residence)<br>OR (TI neighborhood* OR AB<br>neighborhood*)) N2 ((TI safety OR<br>AB safety) OR (TI characteristic*<br>OR AB characteristic*) OR (TI<br>unsaf* OR AB unsaf*))) | Expanders - Apply<br>equivalent subjects<br>Search modes -<br>Boolean/Phrase | Interface -<br>EBSCOhost Research<br>Databases<br>Search<br>Screen -<br>Basic<br>Search<br>Database -<br>APA<br>PsycInfo | Display |
| S2<br>3 | DE "Neighborhoods" | Expanders - Apply<br>equivalent subjects<br>Search modes -<br>Boolean/Phrase | Interface -<br>EBSCOhost Research<br>Databases<br>Search<br>Screen -<br>Basic<br>Search<br>Database -<br>APA<br>PsycInfo | Display |
| S2<br>2 | DE poverty OR (TI poverty OR AB<br>poverty) | Expanders - Apply<br>equivalent subjects<br>Search modes -<br>Boolean/Phrase | Interface -<br>EBSCOhost Research<br>Databases<br>Search<br>Screen -<br>Basic<br>Search<br>Database -<br>APA<br>PsycInfo | Display |

|  |  |  |  |  |
| --- | --- | --- | --- | --- |
| S2<br>1 | (((TI family OR AB family) OR (TI parent* OR AB parent*) OR (TI child* OR AB child*)) N1 (TI relat* OR AB relat*))) | Expanders - Apply equivalent subjects<br>Search modes - Boolean/Phrase | Interface - EBSCOhost Research Databases<br>Search Screen - Basic Search<br>Database - APA PsycInfo | Display |
| S2<br>0 | (DE "family relations") OR (DE "parent-child relations") | Expanders - Apply equivalent subjects<br>Search modes - Boolean/Phrase | Interface - EBSCOhost Research Databases<br>Search Screen - Basic Search<br>Database - APA PsycInfo | Display |

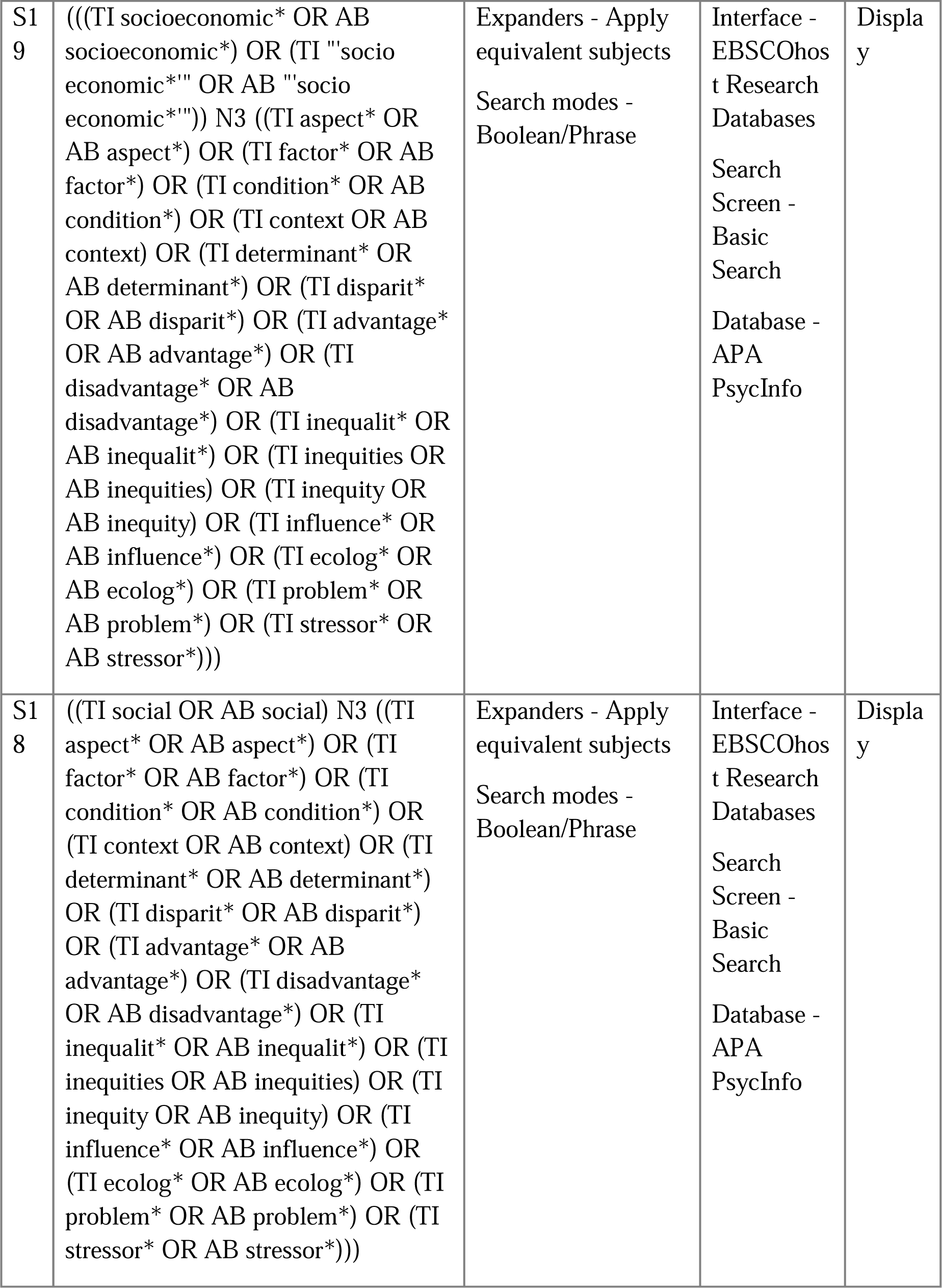

|  |  |  |  |  |
| --- | --- | --- | --- | --- |
| S1<br>7 | DE "Social Ecology" | Expanders - Apply<br>equivalent subjects<br>Search modes -<br>Boolean/Phrase | Interface -<br>EBSCOhost Research<br>Databases<br>Search<br>Screen -<br>Basic<br>Search<br>Database -<br>APA<br>PsycInfo | Display |
| S1<br>6 | DE "social status" OR DE<br>"Socioeconomic Status" | Expanders - Apply<br>equivalent subjects<br>Search modes -<br>Boolean/Phrase | Interface -<br>EBSCOhost Research<br>Databases<br>Search<br>Screen -<br>Basic<br>Search<br>Database -<br>APA<br>PsycInfo | Display |
| S1<br>5 | DE "socioeconomic factors" OR DE<br>"Income Level" OR DE "Lower<br>Class" OR DE "Social Class" | Expanders - Apply<br>equivalent subjects<br>Search modes -<br>Boolean/Phrase | Interface -<br>EBSCOhost Research<br>Databases<br>Search<br>Screen -<br>Basic<br>Search<br>Database -<br>APA<br>PsycInfo | Display |

|  |  |  |  |  |
| --- | --- | --- | --- | --- |
| S1<br>4 | DE "Social Issues" | Expanders - Apply<br>equivalent subjects<br>Search modes -<br>Boolean/Phrase | Interface -<br>EBSCOhost Research<br>Databases<br>Search<br>Screen -<br>Basic<br>Search<br>Database -<br>APA<br>PsycInfo | Display |
| S1<br>3 | DE "Family Socioeconomic Level" | Expanders - Apply<br>equivalent subjects<br>Search modes -<br>Boolean/Phrase | Interface -<br>EBSCOhost Research<br>Databases<br>Search<br>Screen -<br>Basic<br>Search<br>Database -<br>APA<br>PsycInfo | Display |
| S1<br>2 | DE "Social Exclusion" | Expanders - Apply<br>equivalent subjects<br>Search modes -<br>Boolean/Phrase | Interface -<br>EBSCOhost Research<br>Databases<br>Search<br>Screen -<br>Basic<br>Search<br>Database -<br>APA<br>PsycInfo | Display |

|  |  |  |  |  |
| --- | --- | --- | --- | --- |
| S11 | DE "Social Perception" | Expanders - Apply equivalent subjects<br>Search modes - Boolean/Phrase | Interface - EBSCOhost Research Databases<br>Search Screen - Basic Search<br>Database - APA PsycInfo | Display |
| S10 | DE "Socioeconomic Status" | Expanders - Apply equivalent subjects<br>Search modes - Boolean/Phrase | Interface - EBSCOhost Research Databases<br>Search Screen - Basic Search<br>Database - APA PsycInfo | Display |
| S9 | DE "Social Disadvantage" OR DE "Economic Disadvantage" | Expanders - Apply equivalent subjects<br>Search modes - Boolean/Phrase | Interface - EBSCOhost Research Databases<br>Search Screen - Basic Search<br>Database - APA PsycInfo | Display |

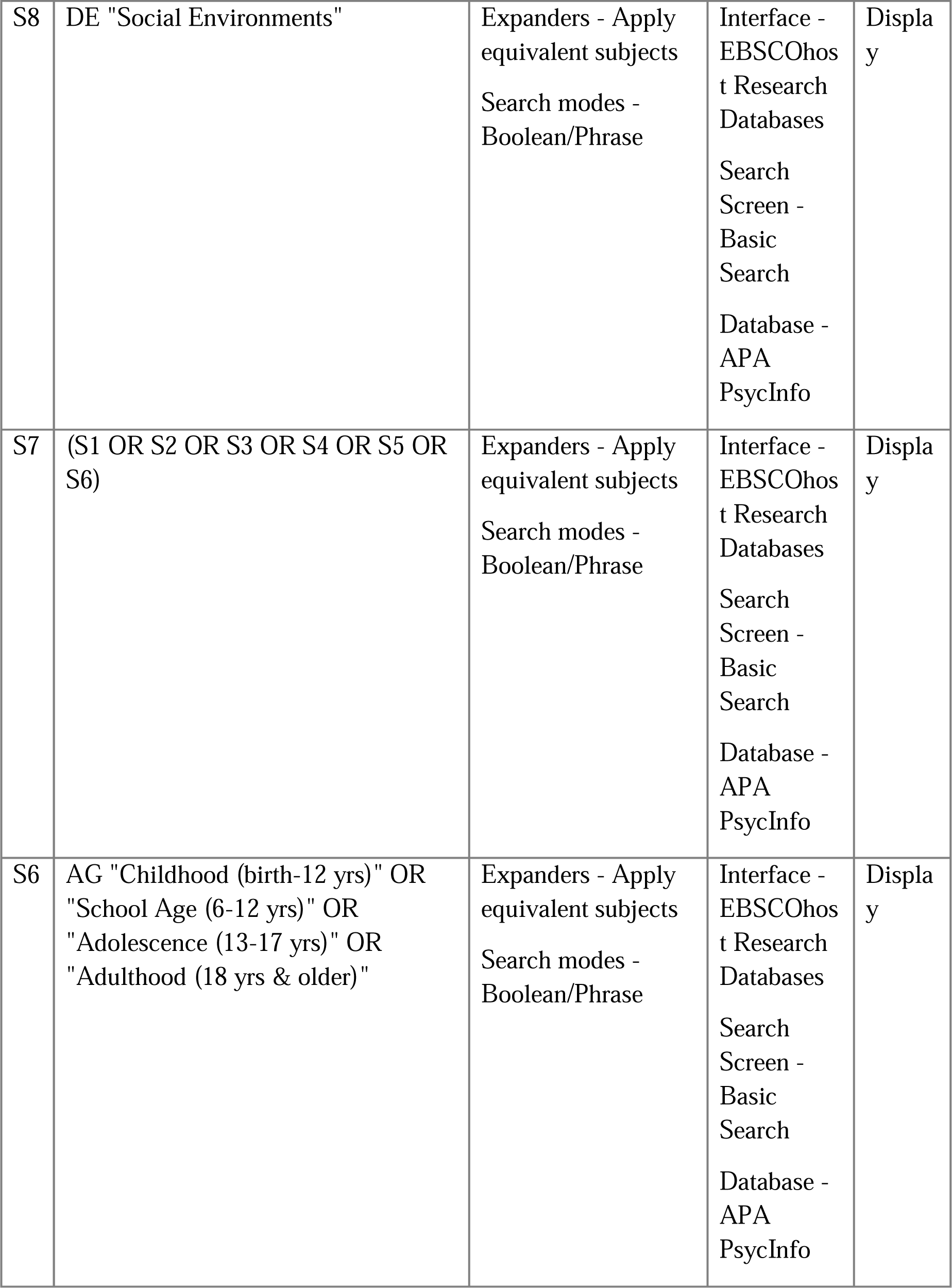

|  |  |  |  |  |
| --- | --- | --- | --- | --- |
| S5 | AB (pediatric* OR paediatric* OR peadiatric* OR toddler* OR baby OR baby* OR babies OR minors OR minors* OR boy OR boys OR boyfriend OR boyhood OR girl* OR kid OR kids OR child OR child* OR children* OR schoolchild* OR adolescen* OR youth* OR teen* OR juvenil* OR underage OR prepubesc* OR pubesc* OR preschool OR school OR school*) | Expanders - Apply equivalent subjects<br>Search modes - Boolean/Phrase | Interface - EBSCOhost Research Databases<br>Search Screen - Basic Search<br>Database - APA PsycInfo | Display |
| S4 | TI (pediatric* OR paediatric* OR peadiatric* OR toddler* OR baby OR baby* OR babies OR minors OR minors* OR boy OR boys OR boyfriend OR boyhood OR girl* OR kid OR kids OR child OR child* OR children* OR schoolchild* OR adolescen* OR youth* OR teen* OR juvenil* OR underage OR prepubesc* OR pubesc* OR preschool OR school OR school*) | Expanders - Apply equivalent subjects<br>Search modes - Boolean/Phrase | Interface - EBSCOhost Research Databases<br>Search Screen - Basic Search<br>Database - APA PsycInfo | Display |
| S3 | SU (adolescent OR youth) | Expanders - Apply equivalent subjects<br>Search modes - Boolean/Phrase | Interface - EBSCOhost Research Databases<br>Search Screen - Basic Search<br>Database - APA PsycInfo | Display |

|  |  |  |  |  |
| --- | --- | --- | --- | --- |
| S2 | SU child | Expanders - Apply equivalent subjects<br>Search modes - Boolean/Phrase | Interface - EBSCOhost Research Databases<br>Search Screen - Basic Search<br>Database - APA PsycInfo | Display |
| S1 | DE pediatrics | Expanders - Apply equivalent subjects<br>Search modes - Boolean/Phrase | Interface - EBSCOhost Research Databases<br>Search Screen - Basic Search<br>Database - APA PsycInfo | Display |

